# Utilizing virus genomic surveillance to predict vaccine effectiveness

**DOI:** 10.1101/2025.06.20.25329795

**Authors:** Jiye Kwon, Ke Li, Joshua L. Warren, Sameer Pandya, Anne M. Hahn, Yale SARS-CoV-2 Genomic Surveillance Initiative, Virginia E. Pitzer, Daniel M. Weinberger, Nathan D. Grubaugh

**Author notes:** Authors listed at the end of the manuscript. These authors contributed equally to this work (Virginia E. Pitzer, Daniel M. Weinberger, Nathan D. Grubaugh).

## Abstract

As new vaccines are being developed for fast-evolving viruses, determining when and how to update them, and what data should inform these decisions, remains a significant challenge. We developed a model to inform these vaccine updates in near real-time and applied it to SARS-CoV-2 by quantifying the relationship between vaccine effectiveness (VE) and genetic distance from mRNA vaccine formulation sequences using 10,156 genomes from Connecticut (April 2021-July 2024) and data from over one million controls, employing a two-stage statistical approach. We showed a strong inverse correlation between spike gene amino acid distance and VE; every 10 amino acid substitutions away from the vaccine sequences resulted in a 15.4% (95% credible intervals (CrI): –2.0%, 34.6%) reduction in VE. Notably, this framework allows us to quantify the anticipated impact of emerging variants on VE, as demonstrated by the predicted 43.4% (95% CrI: –5.7%, 90.1%) drop in VE for the 2023/24 vaccine following the emergence of JN.1 variants based on sequence data alone. By linking amino acid substitutions to VE, this approach leverages genomic surveillance to monitor population-level protection and inform timely vaccine updates.

**Structured Abstract:** 

**Background and aims:** Since the development of the first vaccines targeting the original SARS-CoV-2 virus sequence in 2020, mRNA-based vaccines have been updated three times: targeting Omicron BA.4/BA.5 in 2022, the XBB lineage in 2023, and the KP.2 variant in 2024. While genomic surveillance has advanced our understanding of pathogen diversity, gaps remain in incorporating genomic information to evaluate vaccine effectiveness (VE) against emerging variants. This study aims to characterize the relationship between VE and sequence-based genetic distance, to establish a framework for predicting near real-time changes in the level of vaccine protection from virus surveillance data.

**Methods:** We analyzed 10,156 whole genome sequences of SARS-CoV-2 cases from Connecticut, USA, between April 2021 to July 2024. We first assessed how genetic distance, specifically the number of amino acid substitutions in the spike gene between COVID-19 case sequences and the mRNA vaccine formulation sequence(s), correlates with vaccine protection levels. Incorporating data from over 1 million test-negative controls, we developed a Bayesian time-varying model with autoregressive terms to assess VE at a weekly level. The analysis was adjusted for ZIP-code-level income, age, sex, and prior vaccine doses received. We then employed a random effects meta-regression to explore the relationship between VE and amino acid distance over time. Finally, we used the meta-regression model to estimate potential vaccine protection against emerging variants.

**Results:** We found that spike gene amino acid distance showed a strong negative correlation with VE over time. Stepwise increases in amino acid distance, aligned with sharp VE declines during variant emergence, while accumulation of within-variant changes was also associated with gradual VE decline. Each 10 amino acid increase in distance in the spike gene corresponds to a predicted 15.4% (95% Credible intervals (CrI): –2.0%, 34.6%) reduction in VE. For the 2023/24 updated vaccine, spike distance rose from 12.25 to 30.23, predicting a 43.4% (95% CrI: –5.7%, 90.1%) drop in VE using sequence information alone.

**Conclusion:** Our framework quantifies how the emergence of new variants is expected to affect VE for SARS-CoV-2. By quantifying the relationship between amino acid substitutions and time-varying VE, we leverage intrinsic pathogen features, such as spike amino acid distance, to inform future vaccine updates using genomic sequences. As genomic surveillance data becomes more widely available across pathogens, this framework can serve as a near-real time surveillance tool to infer population-level protection and offers valuable insights for vaccine update decisions.

## Main Text

The antigenic evolution of pathogens, particularly RNA viruses, presents a persistent challenge for disease control and remains a major contributor to global disease burden(*1-3*). As viruses evolve to escape population immunity, they can repeatedly infect individuals, potentially fueling recurrent epidemics(*2, 4*). Viral evolution and epidemiology are deeply interconnected, with transmission both driving and being shaped by viral adaptation(*2, 5*). Achieving population-level immunity through vaccination is a critical public health strategy(*6*). However, when immunity is strain-specific, it leads to opportunities for viral adaptation through immune escape(*7-9*). The resulting decline in population-level vaccine effectiveness (VE) necessitates periodic updates for certain vaccines. Influenza viruses serve as a well-established example, with global surveillance data guiding regular vaccine updates to maintain protection(*10, 11*). More recently, the rapid evolution of SARS-CoV-2 highlights yet another need for frequent vaccine updates. However, COVID-19 vaccine updates have been undertaken annually and largely *ad hoc*. This reflects a broader issue. As vaccines are developed for these and other fast-evolving viruses, determining when and how to update them—and what data should inform these decisions—remains a critical challenge. Addressing this gap is crucial for maintaining VE and optimizing long-term disease control.

In the United States (US), the Advisory Committee on Immunization Practices (ACIP) issues recommendations for the use of updated COVID-19 mRNA vaccines, guided by a rigorous evaluation of VE data in conjunction with clinical, epidemiological, safety, and economic evidence(*12*). The timeline for the 2024/25 COVID-19 vaccine update illustrates this process(*13*). The 2023/24 XBB.1.5 mRNA vaccine became available in September 2023(*14*). In February 2024, ACIP reviewed VE data from the VISION and IVY networks(*15*), supplemented by genomic surveillance on SARS-CoV-2 lineage proportions (e.g., XBB, JN.1) from the Centers for Disease Control and Prevention (CDC)(*15*). In June 2024, ACIP unanimously recommended the 2024/25 KP.2 mRNA vaccine formula for all individuals above 6 months old(*12*); the Food and Drug Administration (FDA) gave full approval to the updated mRNA vaccines from Moderna and Pfizer in August 2024(*16*); and the vaccines became available to the public by October 2024(*17*). However, important gaps remain. Reduced VE of the XBB.1.5 vaccine against the JN.1 was identified in February 2024—months after JN.1 began circulating. Yet, the updated JN.1 lineage-based formula (e.g., KP.2) was not available until October. By this time, KP.3.1.1 had become the dominant lineage, and XEC—a recombinant of two JN.1 lineages—was on the rise(*18*), eroding the benefits of the updated vaccine. This delay underscores the need for a more adaptive vaccine update process. Moreover, proposed FDA guidelines requiring randomized controlled trials to license future COVID-19 updates for use in those under 65 years of age in the US(*19*) further underscore the need for evaluation tools that can assess both when to update the vaccine and the continued effectiveness of previous formulations when updated vaccines are not readily available to all age groups.

Routine genomic surveillance data offer a crucial opportunity to bridge this gap by enabling timely detection of emerging variants with the potential for immune escape. Integrating genomic data with disease surveillance enhances our understanding of infection dynamics, identifies key drivers of change, and informs tailored intervention strategies, including vaccination(*20*). A genomics-informed early warning system hinges on linking four key data sources: virus sequence data, epidemiological data, demographic data, and immunization data(*21*). In this study, we leverage these sources of data to address the need for timely and adaptive vaccine updates. We propose a framework to guide decisions on vaccine updates, specifically for vaccines that stimulate high-affinity neutralizing antibodies against viral antigens. This framework addresses the challenges of rapidly evolving viruses, where antigenic changes in targets of neutralizing antibody can reduce population-level VE. Using COVID-19 data, we present a real-time VE assessment framework that combines data from SARS-CoV-2 genomic surveillance and VE estimates derived using a test-negative design. We achieve this in three main steps: 1) We explore how genetic distance can serve as an early warning signal for reduced VE, by independently analyzing genomic sequences and epidemiological data. 2) We quantify the relationship between spike gene amino acid distance (amino acid substitutions from COVID-19 case sequences relative to the vaccine formulation(s)) and VE, develop a model capable of predicting VE based on amino acid distance, and validate it using a one-month-out prediction framework; and 3) We generate testable predictions of VE in the absence of individual-level vaccination data, demonstrating that changes in vaccine protection can be inferred from genomic sequences alone.

## Results

To investigate how SARS-CoV-2 genetic distance correlates with VE and responds to changes in vaccine formulation over time, we analyzed 10,156 SARS-CoV-2 genomes that we sequenced over 173 weeks through a hospital-based genomic surveillance system (**Figure 1**). Our longitudinal samples captured most of the variants of concern defined as of October 2024, ranging from Alpha to Omicron KP.3.

**Figure 1.**
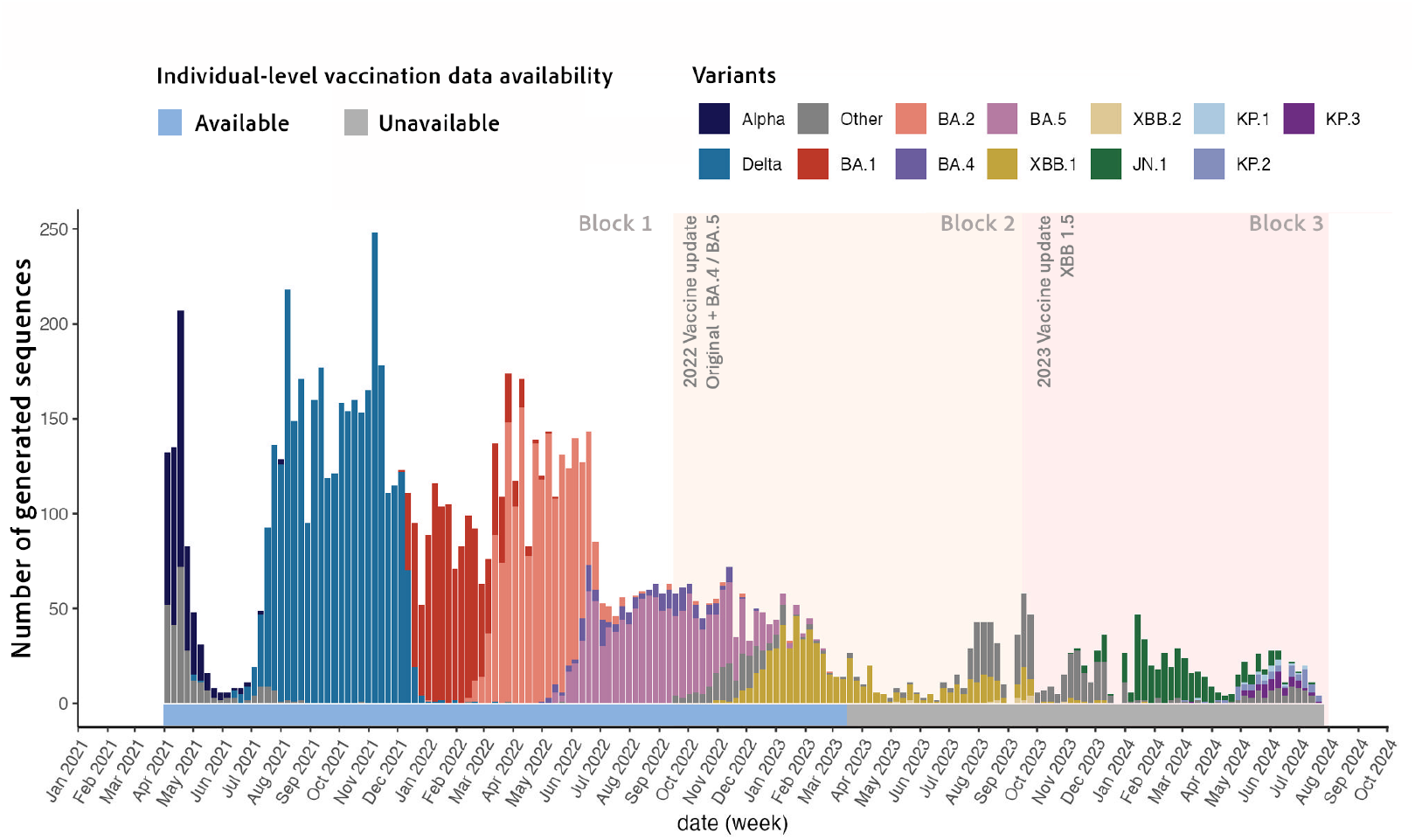
Genomic sequences generated from SARS-CoV-2 cases in Connecticut, US across all vaccine periods (n = 10,156). The number of genomic sequences from SARS-CoV-2 cases is aggregated by week and plotted over the study period, from April 2021 to July 2024. Sequences are colored by different variants of concern (VOCs) prevalent during this period. Background colors represent different vaccine formulation updates: 2020 original monovalent vaccine formulation (white), 2022/2023 bivalent Omicron BA.4/BA.5 formulation (orange), 2023/2024 monovalent XBB 1.5 formulation (pink). The colored bar above x-axis denotes individual-level vaccination data availability: available (light blue) and grey (unavailable).

The study period also captured three COVID-19 vaccine formulations and was divided into three blocks: 1) the original monovalent mRNA vaccine formulation (April 4, 2021 to September 14, 2022); 2) the bivalent (original formula + BA.4/BA.5) mRNA vaccine formulation (September 15, 2022 to September 14, 2023); and 3) the monovalent XBB.1.5 mRNA vaccine formulation (September 15, 2023 to July 27, 2024).

### Genetic distance as an early warning signal during the original vaccine formulation period

To evaluate whether SARS-CoV-2 genetic distance can serve as an early warning signal for declining VE at population level, we first investigated both longitudinal genomic and epidemiological data separately. We analyzed 7,722 genomes from cases that we sequenced during the original vaccine formulation period (i.e., block 1) and examined weekly changes across three key aspects (**Figure 2**).

**Figure 2.**
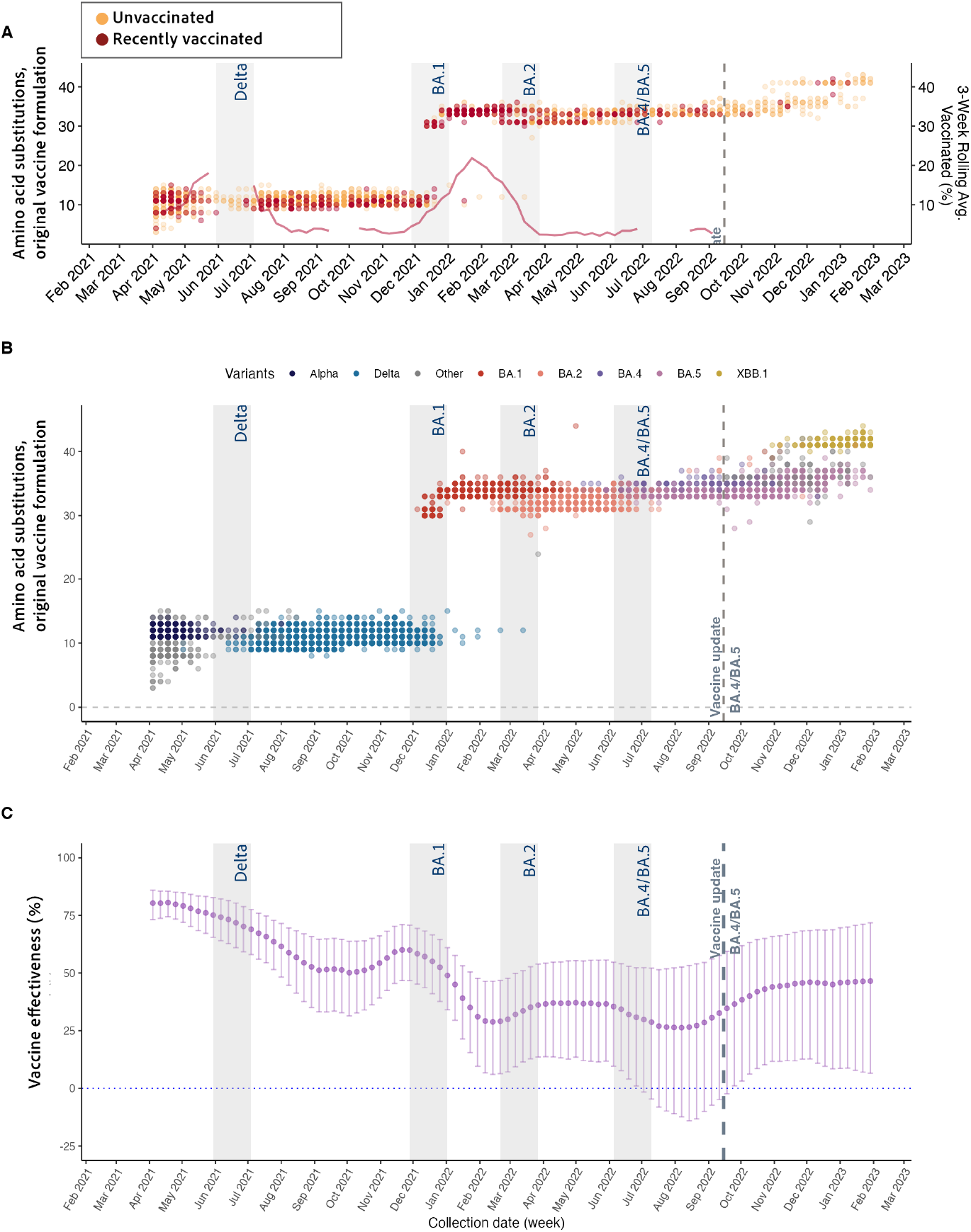
Relationship between spike gene amino acid distance to original vaccine formulation and time-varying vaccine effectiveness for case sequences included in the model calibration, April 2021 to September 2022. Panels (A) and (B) show the number of amino acid substitutions in the spike gene from the case sequences relative to the original formulation of mRNA COVID-19 vaccines. (A) Comparison of sequences between unvaccinated (yellow) and recently vaccinated individuals (red), defined as 14-90 days since the most recent dose. The red line represents the 3-week rolling average of (%) recently vaccinated individuals among sequenced infections over time. (B) Sequences are colored by different variants of concern prevalent during this period (Alpha, black; Delta, blue; Omicron BA.1, red; BA.2, orange; BA.4, purple; BA.5, magenta; XBB.1.5, yellow). (C) Week-level vaccine effectiveness (VE) estimates with 95% credible interval from the time-varying VE model for the period 14-90 days since the most recent dose. Each vertical grey box represents a distinct emergence period, while the dotted line indicates the introduction of the updated bivalent vaccine formulation.

First, we investigated shifts in the proportion of vaccinated individuals among sequenced infections over time (**Figure 2A**). We defined vaccine breakthrough cases as individuals who had received at least one vaccine dose 14 or more days prior to a laboratory-confirmed SARS-CoV-2 infection. From mid-January to mid-February 2022, we observed a rapid rise in breakthrough cases among recently vaccinated individuals (i.e., 14-90 days post-last dose) during the Omicron BA.1 period, comprising roughly 20% of all sequences collected weekly (**Figure 2A**).

Second, we analyzed the weekly average spike gene amino acid distance between the sequenced cases and the vaccine formulation sequence (**Figure 2B**). During the original vaccine formulation period, we showed that the weekly average genetic distance between case sequences and the vaccine sequence ranged from 10 to 34 amino acid substitutions in the spike gene. During the Delta emergence period, the amino acid distance showed little change compared to earlier variants. However, during the Omicron BA.1 emergence period, we observed that average weekly amino acid distance increased from 10.9 in the first week of December 2021 to 33.3 in the first week of January 2022. The amino acid distance remained relatively stable during the BA.2 and BA.4/BA.5 periods. Within-group variations were also evident; BA.1 had the highest within-group variation (mean amino acid distance = 33.55 ± 1.25 standard deviation (SD)) followed by Delta (10.78 ± 1.20 SD). Our analysis of the full spike gene captured a broader range of variations (**Supplemental Figure 2**), offering a more comprehensive view than receptor binding domain alone.

Third, we examined temporal changes in population-level VE (**Figure 2C**). Time-varying VE estimates for the period 14-90 days since the most recent dose gradually declined from 80% (95% credible interval (CrI): 74%, 85%) in April 2021 to around 60% spanning the 6-month period before BA.1 emergence. Following the emergence of BA.1, VE dropped sharply from 58% (95% CrI: 47%, 68%) in early December 2021 to 31% (95% CrI: 13%, 46%) by late January 2022. These low VE estimates persisted during the BA.2 and BA.4/BA.5 periods. We also showed that after bivalent boosters were introduced in mid-September 2022, which reduced the spike amino acid distance between circulating variants and the vaccine formulation sequence (**Supplemental Figure 3**), VE rebounded. In early October, within three weeks of the bivalent booster introduction, mean VE increased by 12 percentage points to 40% (95% CrI: 6.22%, 63.19%), compared to 28% (95% CrI: -9.98%, 54.58%) in late August 2022.

### Quantifying the relationship between amino acid distance and vaccine effectiveness

Based on the strong inverse relationship between the number of amino acid substitutions and VE over time (**Figure 2)**, we hypothesized that population-level amino acid distance in the spike gene can serve as a proxy for vaccine protection. We therefore quantified their direct relationship using our meta-regression model and found that a mean distance of 10 amino acids in the spike gene corresponds to a 15.4% (95% CrI: -2.00%, 34.6%) decrease in predicted VE.

To validate the model’s ability to predict VE using weekly amino acid distance data, we generated one-month-out predictions and compared them to observed VE values (**Figure 3**). We found that the model closely followed the observed VE trends across all prediction months (mean absolute error (MAE) = 3.33) and successfully captured changes surrounding the bivalent vaccine update. All VE values during the eight-month prediction window—four months before and after the vaccine update—fell within the 95% prediction intervals (**Figure 3A**). We also showed that this model outperformed a reference model that had the same autoregressive structure but excluded amino acid distance information (**Figure 3B**); the baseline AR model yielded a higher MAE (MAE = 5.86) and a higher continuous ranked probability score throughout most of prediction months (**Supplemental Figure 4)**. Additionally, the baseline model produced markedly wider prediction intervals, reflecting increased uncertainty in its prediction. These findings highlight the added value of incorporating genomic information in enhancing the model’s accuracy in predicting future VE compared to a model relying solely on historical VE data.

**Figure 3.**
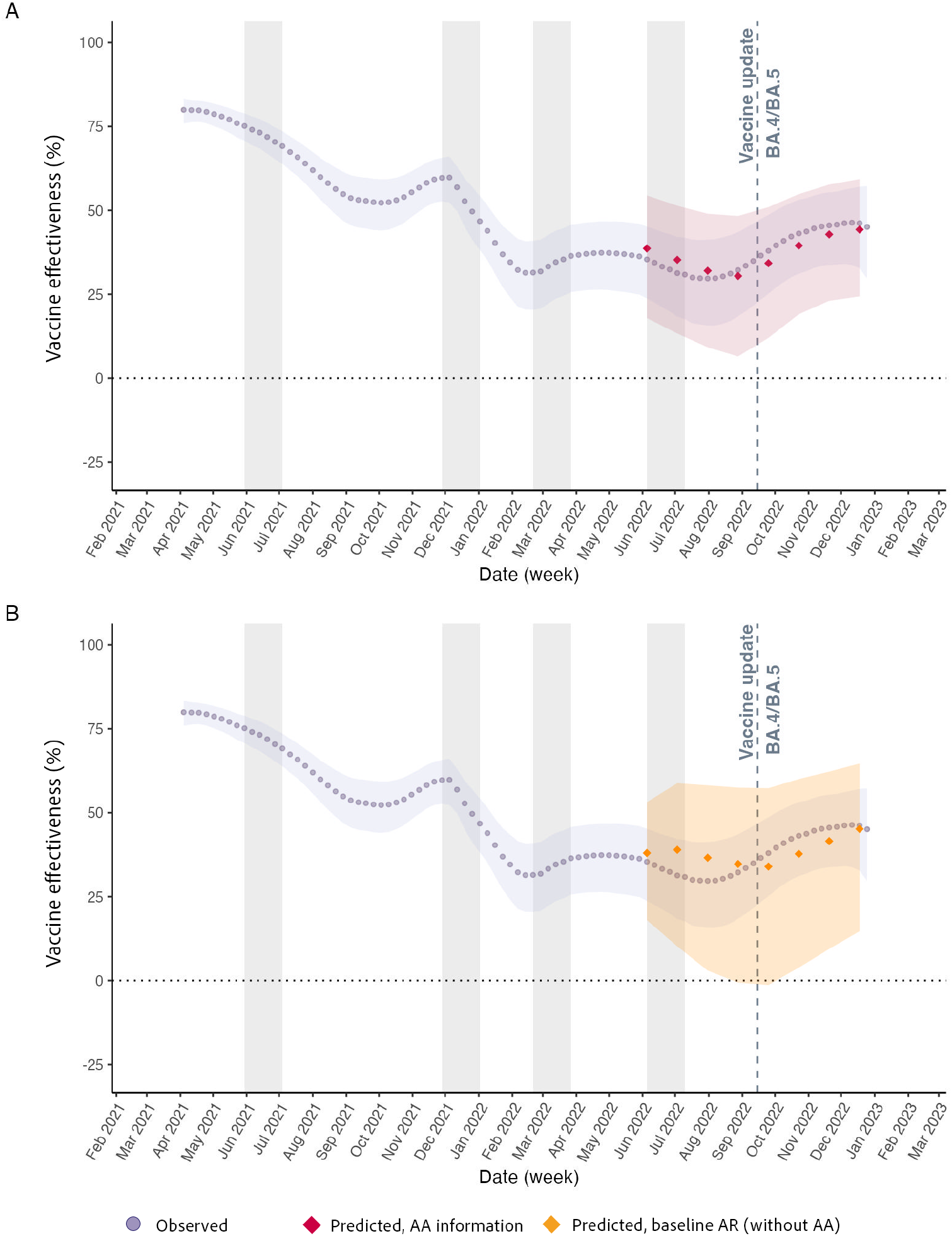
Model validation comparing observed and predicted vaccine effectiveness (VE). The figure compares observed VE from the time-varying model (purple circles) to one-month-ahead predicted VE from (A) the amino-acid-distance meta-regression model (red diamonds), which uses week-level amino acid distance as input, and (**B**) the baseline meta-regression model (orange diamonds), which does not incorporate amino acid distance information. Colored ribbons represent 95% credible intervals for observed (purple) and predicted (red and orange) values.

### Estimating changes in vaccine effectiveness for the XBB.1.5 vaccine formulation: simulating real-world scenarios without vaccination data

After calibrating and validating our model (**Figure 3**), we then sought to translate these findings into a real-world framework to inform changes in predicted vaccine protection as new variants emerged. To evaluate how population-level amino acid distance from the sequence of different COVID-19 vaccine formulations changed across multiple vaccine updates, we extended our model across the different formulation periods. Throughout vaccine update blocks, we utilized only measured weekly amino acid distance as the input to predict the percent reduction in VE; we present VE estimates based on a 3-week rolling average of amino acid distance to enhance visualization.

During the second vaccine (block 2), the increase in minimum amino acid distance to either formulation was more nuanced than in block 1. Block 2 was characterized by the prolonged co-existence of BA.5 and XBB strains, which led to a gradual rather than step-wise increase in population-level mean amino acid distance (**Supplemental Figure 3**). For instance, with the emergence of XBB strains, the mean distance rose from 12.2 in late-December to 18.2 in late-January and 23.7 by mid-September when the 2023/24 XBB vaccine (i.e., third vaccine) became available, marking the first annual COVID-19 vaccine update. This increase in amino acid distance predicted a 34.3% (95% CrI: -2.4%, 70.4%) reduction in VE against the second vaccine (2022/23 vaccine strains) by mid-September.

For the third vaccine update (block 3), we observed a significant increase in mean amino acid distance, mirroring the pattern seen during the original vaccine formulation period (**Figure 4**). Specifically, the mean amino acid distance rose from 12.3 in mid-December 2023 to 30.2 in mid-January 2024, corresponding to the transition from XBB to JN.1 dominance. Based on this increase in amino acid distance, we predicted a 43.4% (95% CrI: -5.7%, 90.1%) reduction in VE by mid-January 2024. Our findings also suggest a further 9.8% decline in VE against circulating variants by June 2024, coinciding with the emergence of KP lineages, reaching an estimated 53.2% (95% CrI: 7.0%, 110.4%) reduction compared to VE against the XBB.1.5 vaccine strain. This approach demonstrates how the model can support real-time surveillance by quantifying expected declines in vaccine protection as new variants emerge.

**Figure 4.**
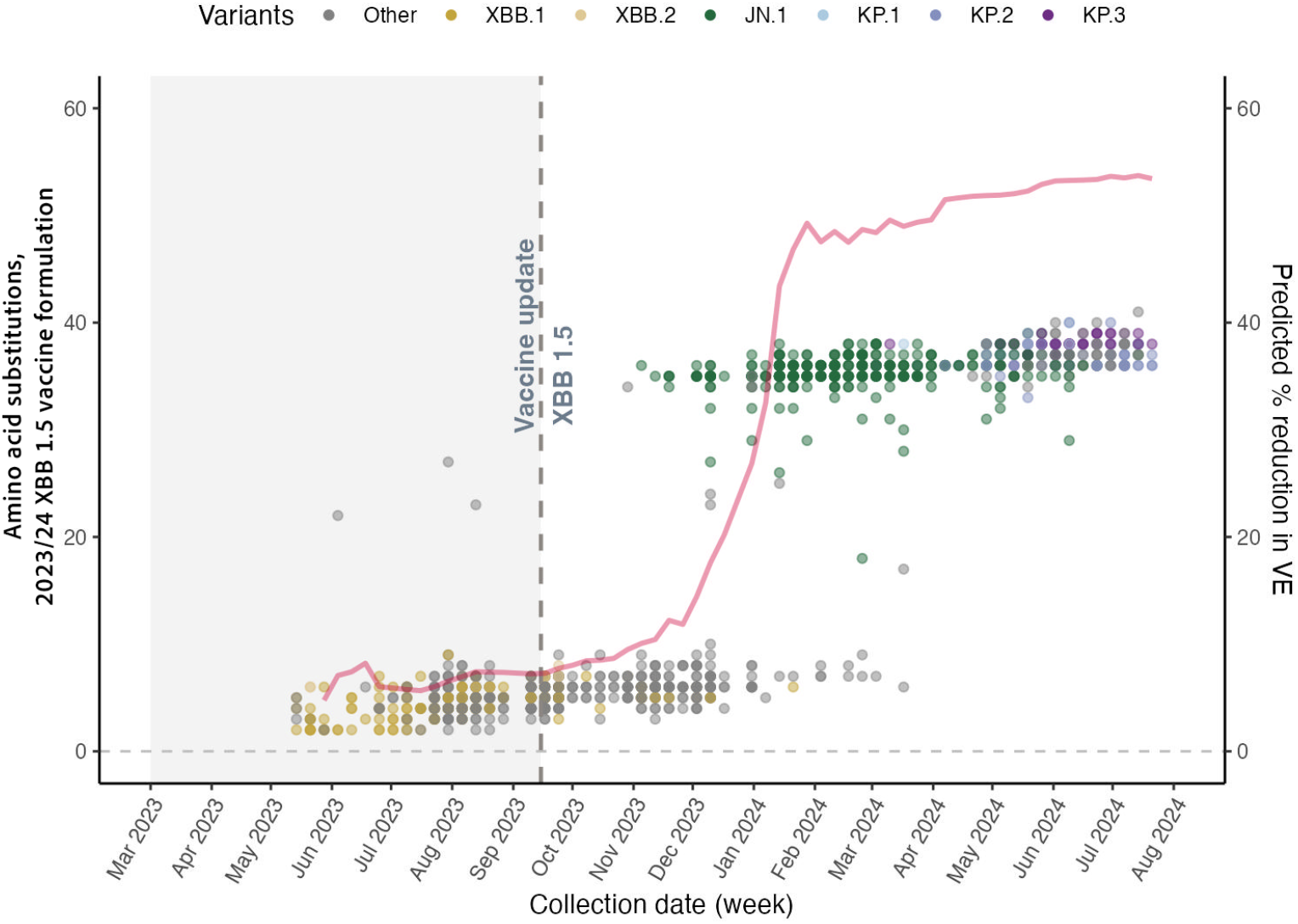
Amino acid differences in spike gene between case strains and the XBB.1.5 formulation of mRNA COVID-19 vaccines. Sequences are aggregated by week from May 2023 to July 2024. The grey dotted line represents the introduction of the XBB.1.5. vaccine update. The grey shaded area represents sequences that were collected prior to XBB.1.5 vaccine availability. Sequences are colored by different variants of concern prevalent during this period. The red line represents the weekly mean predicted percent reduction in vaccine effectiveness (VE) based on the 3-week rolling average of amino acid distance.

## Discussion

Our analysis estimated time-varying COVID-19 VE at the population level and quantified its direct relationship with SARS-CoV-2 genomic data from community cases. The time-varying VE framework that we introduce is useful by itself for identifying and predicting temporal changes in VE. However, complementing the framework with genomic information provides additional precision for generating one-month-ahead predictions of VE. We found that virus genomic data, in the form of amino acid distance from the vaccine formulation sequence, can serve as an early warning to detect shifts in predicted VE and provides insights into the underlying causes of these changes. Combining time-varying VE estimates with the association between VE and amino acid distance allows us to quantify how the emergence of new variants is expected to affect VE.

We demonstrated a clear association between increasing amino acid distance in the SARS-CoV-2 spike gene from the vaccine formulation and declining population-level VE. This aligns with a growing body of research highlighting the critical role of spike mutations on antibody neutralization and immune escape(*22-24*). While many prior works on immune escape have focused on variant-specific analyses(*25*), *in vitro* characterization of specific mutations(*26*), or mapping of viral evolution across variants using antigenic cartography(*27*), our approach offers a complementary population-level perspective. We provide a variant-agnostic and practical metric for real-time monitoring of potential reductions in VE. This contrasts with traditional VE studies that often rely on retrospective analyses of observational data.

For SARS-CoV-2, genetic distance to the vaccine sequence has previously been explored as a correlate of vaccine efficacy based on clinical trial data (*28, 29*) and through predictive models using publicly available data(*30*). Previous models focused on forecasting and characterizing specific mutations of concern(*30*); our study expands the application by directly estimating VE using genomic surveillance data. A key strength of our study lies in the comprehensive surveillance data, which encompasses four years of SARS-CoV-2 genomic surveillance from a single hospital system, uniquely linked to detailed clinical metadata. This integration of epidemiological and genomic data—rather than relying on public databases— allows us to quantify the relationship between VE and temporal changes in amino acid distance, advancing our understanding of vaccine protection in an evolving antigenic landscape. Furthermore, our Bayesian approach to estimating time-varying VE with autoregressive terms enables us to provide robust and informative estimates even for weeks with limited sequence availability.

Vaccine formulation updates are sometimes necessary to provide sufficient protection against emerging immune escape variants(*31, 32*). In the case of SARS-CoV-2, the emergence of antigenically distinct lineages has been characterized by the accumulation of a large number of mutations, which thus far has been difficult to predict(*2*). COVID-19 vaccine formulation updates have generally been implemented annually since 2023. However, the degree of antigenic divergence from the existing vaccine formulation varied among the variants that prompted these updates. For instance, the emergence of Omicron lineages, characterized by over 30 amino acid substitutions in the spike protein alone compared to ancestral lineages, prompted the first vaccine update(*33, 34*). Our analysis showed that an increase in amino acid distance of approximately 30 substitutions following the emergence of Omicron BA.1 corresponded to an estimated 40% decrease in VE. The emergence of JN.1 variants in November-December 2023 was characterized by a similarly large increase in amino acid distance relative to the XBB.1.5 (2023/24 vaccine update) formulation, which we predict should have resulted in a similar ∼40% decrease in VE.

However, the antigenic divergence of XBB.1.5 from the preceding BA.4/BA.5 vaccine (2022/23 update) resulted in comparatively smaller predicted VE reduction, estimated at ∼20% upon emergence in late December 2022 and ∼35% when the updated vaccine was implemented (September 2023). Our framework could help identify when emerging variants are associated with large (vs small) declines in VE, better guiding when COVID-19 vaccine formulation updates are needed, as opposed to the current practice of updating the vaccine annually. This could enable preparedness efforts before clinical data fully accumulate, allowing for a more timely and dynamic response system in the future. The threshold amino acid distance dictating vaccine updates can be adjusted based on desired vaccine protection targets, but the key insight is that waiting for full laboratory and clinical confirmation may not be necessary to initiate a response. Instead, amino acid distance can serve as an effective early indicator, with additional data validating and refining decisions.

Limitations to our study include the lack of disease severity information among outpatients; therefore, our VE estimates are not outcome specific (e.g., hospitalization, death, etc.) within this population. While outcome-specific VE would provide further insights, our focus on outpatient data aligns more closely with the population we aimed to study, namely people seeking healthcare due to COVID-19-related symptoms in the community setting. Inpatient data were excluded due to the potential for bias introduced by routine testing protocols prior to procedures (e.g., surgeries). Similarly, we identified individuals with high frequency of testing (more than 50 times between April 2021 and March 2023) as likely undergoing routine testing. Although we made efforts to flag and account for their potential influence in our models, some residual bias may remain. Such individuals may be more likely to test negative and either more likely to be vaccinated (if they represent very “COVID-aware” individuals) or less likely to be vaccinated (if routine testing was required by their employer as a condition of being unvaccinated). Furthermore, our VE estimates are not vaccine specific. We used the mRNA vaccine sequences for the Pfizer^®^ BNT162b2 vaccine, which were similar for the Moderna^®^ vaccine, to define amino acid distance. While these vaccines may not fully represent all COVID-19 vaccines used in our study population, they account for over 97% of doses administered in the US(*35-37*). Additionally, we did not account for potential bias related to depletion-of-susceptible people, which can occur when natural immunity from infection is acquired faster in the unvaccinated group than in the vaccinated group, potentially underestimating VE and leading to incorrect conclusions about time-varying effectiveness(*38, 39*). Lastly, we implemented a two-stage approximation to a full hierarchical Bayesian model; we addressed first-stage uncertainty through a meta-regression framework with autoregressive random effects, but this remains an approximation. While VE estimates may vary depending on the population and setting, our approach provides a framework that is adaptable to other contexts, provided genomic surveillance data are available and can be linked to patient information for cases and controls.

Uncertainties remain in the seasonality of SARS-CoV-2(*40, 41*) and the optimal frequency of COVID-19 vaccine updates. Our study provides a framework for the early identification of emerging variants associated with declines in vaccine protection, which could allow for more adaptive decision-making on COVID-19 vaccine updates. Currently, committees like the ACIP play a pivotal role in making policy recommendations based on comprehensive review of clinical, epidemiological, safety, and economic data(*12*). Our framework supplements current work as we demonstrate how genomic data can be integrated with more traditional epidemiological data sources to address the need for timely, reliable evidence to guide vaccine updates. It also provides reassurance during periods when the current vaccine is expected to remain effective, strengthening public confidence in VE messaging. Ultimately, vaccine protection is shaped by a complex interplay of factors, including vaccine properties, evolutionary dynamics of the virus, biological factors influencing host immunity, and selective pressure within a population(*40*). This highlights the need for continuous monitoring and robust data. Genomic surveillance is integral to this process, enabling more informed decision-making and effective public health strategies.

## Data Availability

We used the R statistical software (v4.4.1) for all statistical analyses and visualization. Data and code used in this study are publicly available on Github: https://github.com/jiyekwon/Yale_COVIDVE.

## Author contributions

Conceptualization: JLW, VEP, DMW, NDG

Methodology: JK, KL, JLW, VEP, DMW, NDG

Investigation: JK, JLW, KL, AMH, VEP, DMW, NDG

Visualization: JK, JLW, DMW

Funding acquisition: VEP, DMW, NDG

Project administration: VEP, DMW, NDG

Supervision: VEP, DMW, NDG

Writing – original draft: JK

Writing – review & editing: JK, KL, JLW, SP, AMH, VEP, DMW, NDG

