## Supplemental Figure 1, Supplemental Figure 2, Supplemental Figure 3, Supplemental Figure 4 for "Utilizing virus genomic surveillance to predict vaccine effectiveness"

### From Sequences to Real-Time Vaccine Effectiveness Evaluation

**The PDF file includes:**

Materials and Methods
Figs. S1 to S4
References (42-59)

### Materials and Methods

Ethics statement

The study protocols for pathogen genomic sequencing of remnant diagnostic samples (IRB Protocol ID: 2000033281) and impact of SARS-CoV-2 genetic variants on immune escape and vaccine breakthroughs (IRB Protocol ID: 2000031374) was reviewed and approved by the Yale University institutional review board.

Data sources and genetic sequences

We used whole genome sequencing (WGS) data collected from the Yale New Haven Hospital (YNHH) system in Connecticut (CT), US between the week of 4$^{th}$ April 2021 to the week of 21$^{st}$ July 2024 through the Yale SARS-CoV-2 Genomic Surveillance Initiative. Sample collection, RT-qPCR, and sequencing protocols have been described in detail elsewhere(*42, 43*). Metadata and vaccination records were obtained from the YNHH system and Center for Outcomes Research and Evaluation and matched to WGS data using unique sample identifiers. We used standardized ZIP-code-level CT income data as a potential covariate, with raw data obtained from the US Census Bureau(*44*).

We highlighted emergence periods for better visualization and understanding of variant timelines. For uniformity, we applied the same emergence period definitions previously established in Chen et al.(*43*) In short, these periods represent short intervals during which two variants co-circulated, defined as the time from when a variant accounted for 5% of Global Initiative on Sharing All Influenza Data (GISAID)-reported cases in CT to when it reached its maximum frequency in the population(*43*).

Epidemiological data

Our study focused on CT residents to ensure consistency in surveillance and vaccination policies. Cases were defined as individuals who tested positive for SARS-CoV-2 infection via PCR and who had samples sequenced at the WGS level. To maintain a consistent definition of VE, only samples taken from outpatient settings were included. We excluded samples for whom WGS information was not available or did not meet the required genome sequence coverage (i.e., genome coverage >95%). Other exclusion criteria included duplicate samples from an individual (to exclude chronic infections) and samples from non-CT residents. Given the vaccine rollout and eligibility timelines differed across age groups, we only included adult patients (18+

years) at the time of sample collection. Cases were aggregated at the week level to reduce sparsity, and subsequent analyses were conducted at the week level.

To assess VE over the first 103 weeks, corresponding to the first two years of the data collection (April 4, 2021 to March 19, 2023), we obtained individual-level metadata from test-negative controls. Controls were selected from samples taken from individuals in outpatient settings who met all criteria for the YNHH system testing initiative, but who tested negative for SARS-CoV-2 infection via PCR. If an individual had both negative and positive test results, we only included the positive test as a case. Any subsequent negative test from the same individual within two weeks of a positive test was excluded to avoid potential misclassification.

A patient's vaccination status was determined at the time of sample collection, and time since last vaccination was calculated as the difference between the sample collection date and date of last vaccine receipt. Primary analyses were conducted based on time since last vaccination to account for waning immunity, and uniform criteria were applied regardless of the vaccine manufacturer or number of doses received. Time since last vaccination was categorized as: <14 days, 14-90 days, 91-180 days, ≥181days. The total number of doses received more than 14 days prior to sample collection was also counted for each individual and included as a covariate in our model. For visualization, we plotted 3-week rolling averages to show the proportion of recently vaccinated individuals among both the cases with collected sequences and test-negative controls over time. This allows us to observe trends in vaccine coverage in both groups.

To ensure that our controls were representative of the population, vaccine coverage among controls was compared against available population vaccine coverage data in CT **(Supplemental Figure 1)**. We obtained data on the CT vaccination trends from the CDC(*45*).

Vaccine updates and corresponding sequences

Comirnaty® (Pfizer-BioNTech) or Spikevax® (Moderna Tx.) were the most widely available and used COVID-19 vaccines in the US. Updates to these two mRNA-based vaccines mostly followed a similar timeline throughout the study period with three updates as of October 2024(*46*).

Vaccine blocks were defined based on FDA approval dates, though the exact cut-off dates were selected arbitrarily for analysis purposes. The FDA authorized the use of the bivalent (original formula + BA.4 / BA.5) vaccine as a booster dose on August 31, 2022(*47*); we assigned

September 15, 2022 as the start of bivalent vaccine availability, regardless of the vaccine manufacturer(*48*). The monovalent XBB.1.5 formulation was approved on September 11, 2023(*49*); we assigned September 15, 2023 as the beginning of the third block.

Genetic distance model

We defined genetic distance as the number of amino acid substitutions between the case and mRNA vaccine formulation spike gene sequences. To extract the spike gene region from aligned WGS, we used `bedtools`(*50*) to trim the specified genomic coordinates at the nucleotide level. We then translated the nucleotide sequences to amino acid. We measured genetic distance using Hamming distance, which calculates the number of mismatched positions between aligned sequences(*51*). Hamming distance was selected as it has been shown to correlate well with antigenic distances and has been effectively used in influenza vaccine research(*51-53*).

For each vaccine block, we compared case sequences to the corresponding vaccine sequence available during that time period. For example, sequences from block 1 were compared against the original mRNA vaccine sequence, while sequences from block 3 were compared against the 2023/2024 XBB.1.5 formulation. For block 2, we used the minimum amino acid differences between the case strain and either of the two strains in the bivalent vaccine.

Time-varying vaccine effectiveness (VE) model

We assessed time-varying VE using a hierarchical Bayesian time-series method, building on the widely used test-negative design that compares the prevalence of vaccination among SARS-CoV-2 cases and test-negative controls. The binary outcome variable for an individual *i* at time *t* ($Y_{it}$) is equal to one if the individual is a case and zero if they are a control. We used logistic regression to model the probability of being a case or control for each individual and time point ($p_{it}$) as a function of vaccination status and other covariates. Additionally, we extended the conventional test-negative design approach by incorporating correlated random effects to allow the association between vaccination status and case/control probability to smoothly change across time. Specifically, the model is given as:

$$Y_{it}|p_{it} \sim \text{Bernoulli}(p_{it}), \text{logit}(p_{it}) = \beta_{0t} + \sum_{j=1}^{c} \beta_{jt} \mathrm{Z}_{ijt} + \mathbf{x}_i^{\mathrm{T}} \boldsymbol{\eta}$$

**( 1 )**

where $\beta_{0t}$ is the time-varying intercept parameter that accounts for changes in baseline risk of SARS-CoV-2 infection for each week *t*; $Z_{ijt}$ is an indicator variable for the vaccination category (0 (reference): unvaccinated, 1: <14 days, 2: 14 – 90 days, 3: 91 days – 6 months, 4: >6 months) of individual *i* in week *t*; $\beta_{jt}$ is the time-varying regression parameter that describes the association between vaccination category *j* and the probability of being a case; and $\mathbf{x}_i$ is a vector of covariates for individual *i* for which the corresponding effect estimates do not change across time. The vector of other covariates includes age group, standardized income at the zip-code level, gender, number of vaccine doses received before sample collection, and a "routine flag" (binary variable) to indicate individuals who are thought to be testing routinely (defined as 1 for individuals with more than 50 testing records over the 103 weeks included in our dataset and 0 otherwise).

The time-varying parameters, $\beta_{0t}$ and $\beta_{jt}$, were modeled as a function of a "global" fixed effect parameter and independent (across parameter) autoregressive correlated random effects, such that:

$$\beta_{jt} = \left(\beta_j + \theta_{jt}\right);\ \theta_{jt} |\ \theta_{jt-1}, \rho_j, \tau_j^2 \sim \mathrm{N}\left(\rho_j \theta_{j,t-1}, \tau_j^2\right), j = 0, \dots, c.$$

The autoregressive structure specified for the $\theta_{jt}$ parameters allows both intercept and coefficients to evolve dynamically while borrowing strength across nearby time points. We allow the data to determine the strength of temporal correlation by estimating the $\rho_j$ parameters. To complete the model structure, we assigned prior distributions to the remaining model parameters. Specifically, we assigned independent log-gamma(3, 2) priors to the precision parameters $\left(\frac{1}{\tau_j^2}\right)$ of the random effects. For the fixed effects, including $\boldsymbol{\eta}$ and $\beta_j$, we used weakly informative Normal(0, $10^3$) priors. Additionally, we selected independent Uniform(-1, 1) distributions for the autoregressive parameters ($\rho_j$).

We used R-INLA (integrated nested Laplace approximations) (www.r-inla.org)(*54, 55*) to fit the model. R-INLA is a computationally efficient approximation to a widely used Markov chain Monte Carlo (MCMC) approach, and provides marginal posterior inference for the included model parameters(*56*). We generated 10,000 samples from an approximated marginal posterior distribution for each parameter in parallel using `pbmcapply` and `parallel` R packages. Along with posterior means of $\beta_{jt}$, the quantile based, equal tailed 95% credible intervals (CrI) of $\beta_{jt}$ were constructed for each vaccination category. For better visualization, both the posterior mean and CrI for 1- $\exp(\beta_{jt})$ were also calculated and plotted, representing VE (%) at each week.

<u>Quantifying the relationship between amino acid distance and vaccine effectiveness</u>

We used a random effects meta-regression model fitted in the Bayesian setting to quantify the relationship between amino acid distance and VE over time, focusing on the 14-90 days post-vaccination category (*j = 2*) to represent short-term vaccine-induced immunity. We modeled the outcome variable, the observed log OR of infection at time *t* ($\hat{\beta}_{2t}$) from the time-varying VE model as following a normal distribution centered around the latent true log OR ($\beta_{2t}$), with variance defined by the squared posterior standard deviations ($\hat{\sigma}_t{}^2$). Then, the true but unobserved log OR is modeled as a function of amino acid distance and random effects. Specifically, the full model is given as:

$$\hat{\beta}_{2t}|\beta_{2t} \sim \mathrm{N}\left(\beta_{2t}, \hat{\sigma}_t{}^2\right);\ \beta_{2t} = \gamma_0 + \gamma_1 AA_t + \alpha_t + \varepsilon_t$$

where $\gamma_1$ is the key regression parameter that quantifies the association between amino acid distance and the relative risk of infection; $\gamma_0$ is the baseline intercept that accounts for variations in baseline risk across all time points; $\alpha_t$ captures time-varying random effects with an autoregressive structure similar to the first-stage model; and $\varepsilon_t$ is an independent error term that follows a zero-mean normal distribution. To complete the model structure, we assigned weakly informative priors to the remaining parameters. Specifically, we assigned Normal(0, $10^3$) priors for the fixed effects, including $\gamma_0$ and $\gamma_1$. For the random effects ($\alpha_t$), we assigned a Uniform(-1, 1) prior to the autoregressive parameter ($\rho_A$), consistent with our first-stage model, and a Gamma(0.01, 0.01) prior to the precision parameter governing the variability.

We used MCMC methods to fit the model and sampled from the joint posterior distribution of all model parameters using the `rjags` package(*57*) in R. We initialized four MCMC chains with an adaptive phase of 500,000 iterations, followed by 100,000 iterations after convergence. Convergence was assessed via Geweke diagnostics, which yielded Z-scores of -0.648 for $\gamma_0$ and 0.675 for $\gamma_1$, indicating no obvious signs of non-convergence. Posterior inference was based on posterior means and a 95% CrI for key parameters including $\gamma_1$.

### Model validation and prediction

We assessed the predictive performance of the random effects meta-regression model using a one-month-out validation framework. This approach was designed to simulate real-world scenarios where genomic surveillance data are available, but individual-level patient metadata are limited. In such contexts, amino acid distance data serve as the primary input for predicting VE, which is derived from estimates of $\beta_{2t}$ from the meta-regression model.

We generated predictions over an eight-month time period from June 2022 to January 2023, spanning four months before and after the bivalent vaccine update. Specifically, for each held-out month $k$, we excluded the observed $\hat{\beta}_{2t}$ values from the model and used only amino acid distance data for that month as input. The observed $\hat{\beta}_{2t}$ values from months 1 to $k$-1 were retained in the model to inform the posterior distribution of the time-varying parameters. The model was used to predict $\beta_{2t}$ for month $k$, given the amino acid distance data for that month. For example, to predict the $\beta_{2t}$ for June 2022, the model was given $\hat{\beta}_{2t}$ data up to May 2022 and was asked to predict the $\beta_{2t}$ for June 2022 based on the amino acid distance data for June 2022. This process was repeated for each month in the validation period.

We fit the meta-regression model described in the previous section separately for each leave-out month using MCMC methods. We implemented the model using four MCMC chains with 50,000 iterations followed by 10,000 iterations after convergence. The model's predicted $\beta_{2t}$ values were compared to the observed $\hat{\beta}_{2t}$ to assess its predictive performance. The MAE was used as the primary metric to quantify prediction error. A baseline autoregressive model, using a similar one-month-out validation framework and MCMC settings, was also evaluated; this model did not incorporate amino acid distance information, instead predicting VE based only on the autoregressive terms.

To compare the predictive performance of these two models, we calculated CRPS. Unlike the MAE, which compares point predictions to observed values, the CRPS is a generalized MAE that evaluates the entire predictive distribution(*58, 59*). We computed CRPS for each one-month-out prediction for both models to provide a more comprehensive assessment of their probabilistic forecasting capabilities.

### Data availability

All genome sequences used for this analysis and a subset of the associated metadata (accession number, virus name, collection date, originating lab and submitting lab, and the list of authors) in this dataset are published in GISAID's EpiCoV database. Vaccination metadata are provided in our GitHub repository.

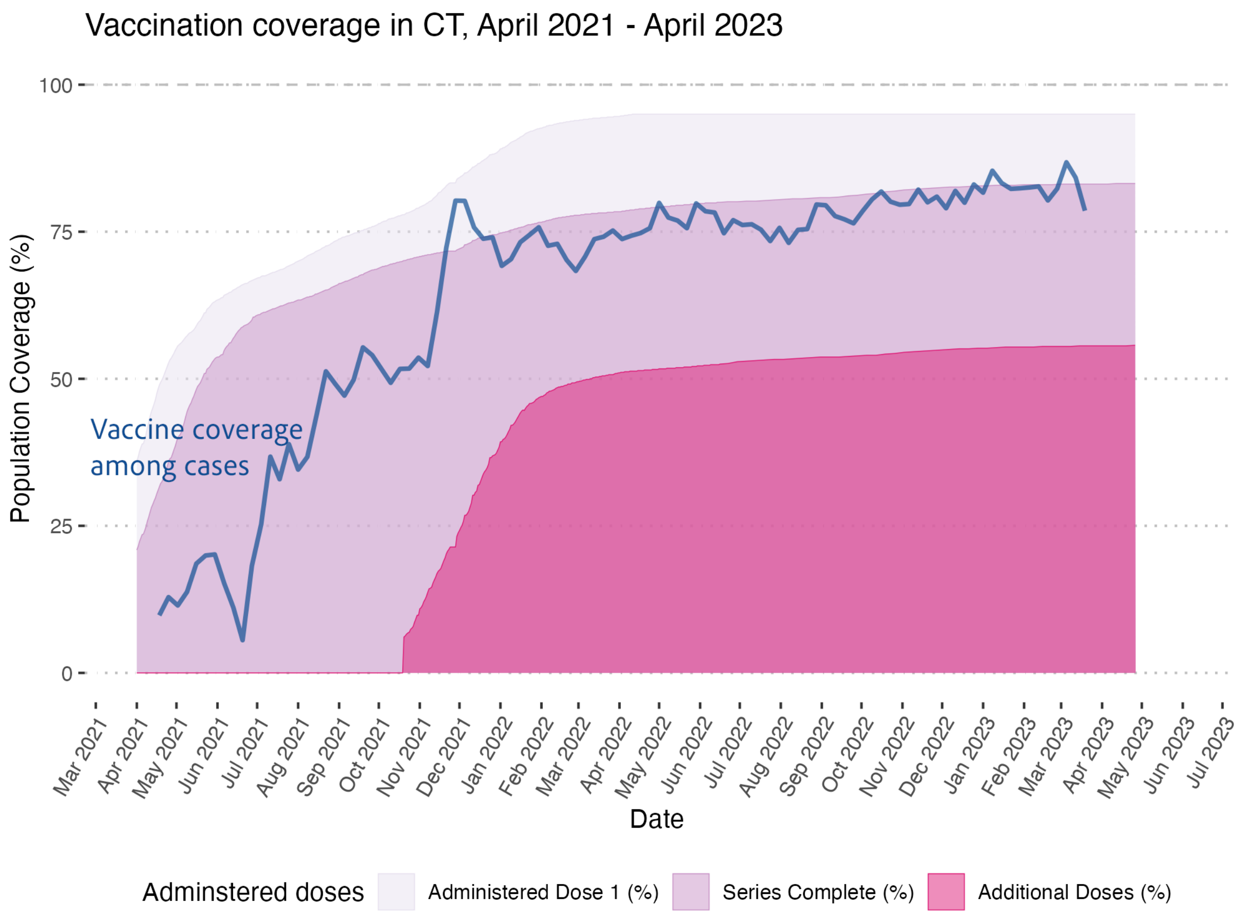


**Fig. S1. Vaccine coverage trends in Connecticut (April 2021 – May 2023).**
Weekly vaccine coverage data for Connecticut, USA, based on publicly available CDC data. Colors indicate different administered dose categories: dose 1, primary series completion, and additional doses. Coverage is presented as the percentage of the population receiving each dose type over time. Coverage among cases (blue line) is presented as 3-week rolling average.

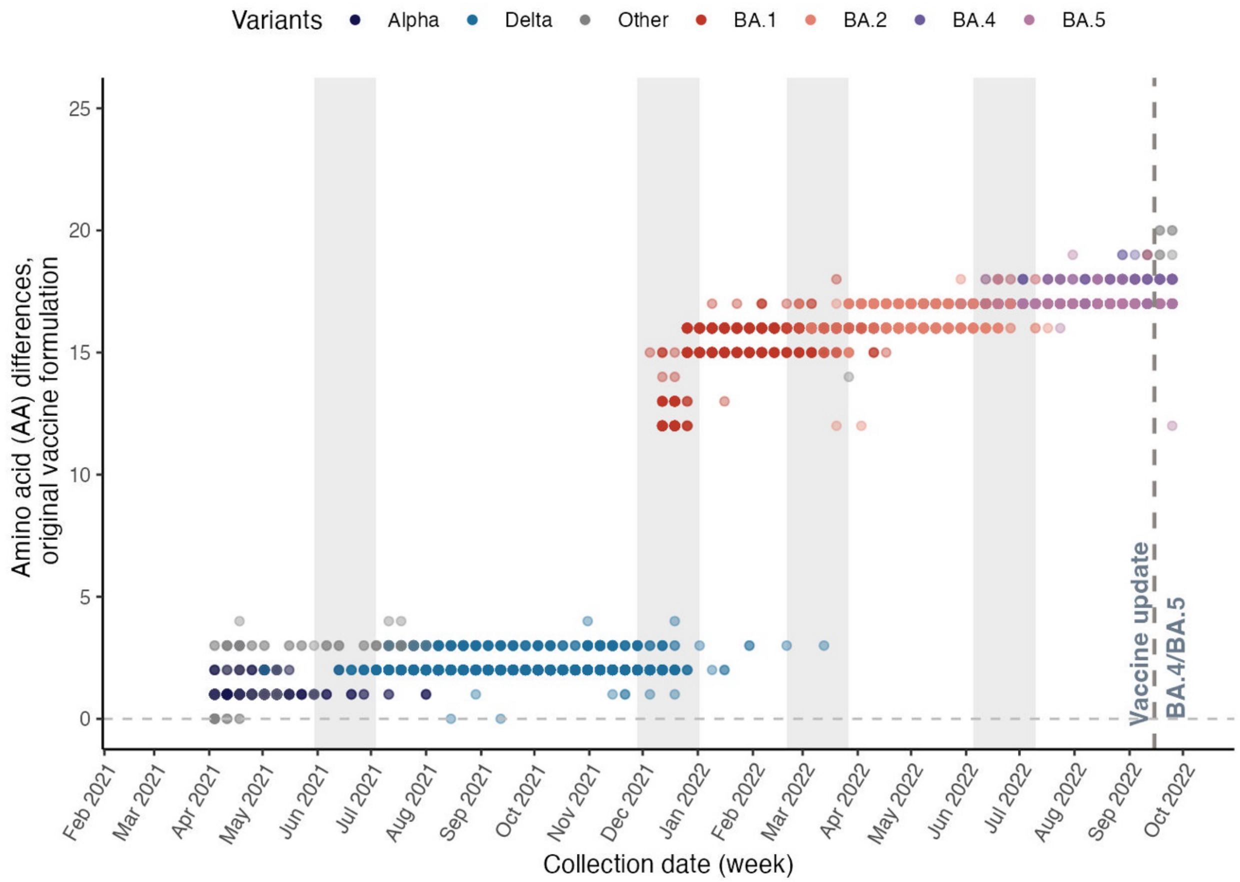


**Fig. S2. Amino acid differences in the receptor binding domain relative to the original mRNA COVID-19 vaccine strain, April 2021 to September 2022.** Figure shows weekly aggregated amino acid differences in the receptor binding domain region between the case strain and original formulation of mRNA COVID-19 vaccines. Sequences are colored by the predominant variants of concern prevalent during this period (Alpha, black; Delta, blue; Omicron BA.1, red; BA.2, orange; BA.4, purple; BA.5, magenta).

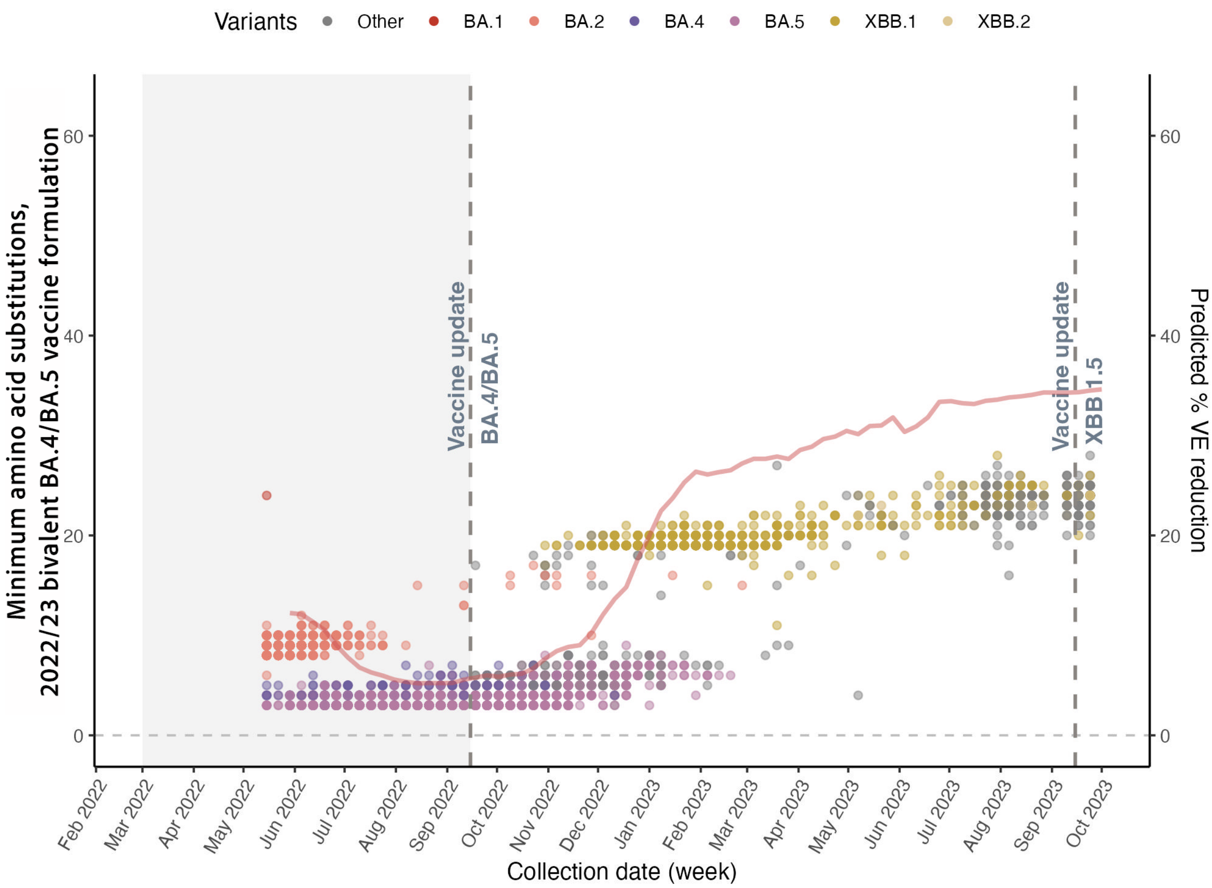


**Fig. S3. Amino acid differences in the SARS-CoV-2 spike gene relative to the bivalent mRNA COVID-19 vaccine strains, May 2022 to September 2023.** Figure shows the weekly aggregated minimum number of amino acid substitutions in the spike gene between SARS-CoV-2 case sequences and either of the two strains included in the bivalent mRNA COVID-19 vaccine formulation. Sequences are colored by the predominant variants of concern circulating during this period.

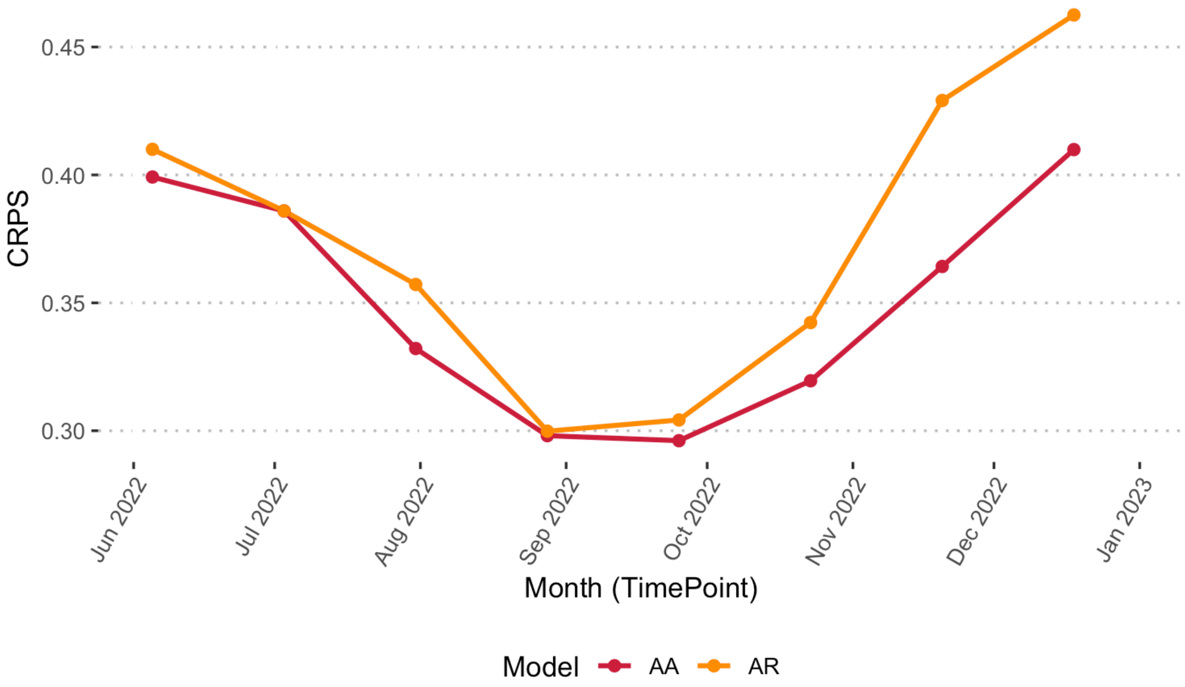


**Fig. S4. Continuous ranked probability score (CRPS) model comparison.** Figure displays the CRPS values for the amino acid distance informed model (red line) and the baseline autoregressive (AR) model (orange) over the eight-month prediction and validation period. Lower CRPS values indicate better performance.
